# Associations Between Salivary Cortisol, DHEA-S, and Alpha-Amylase and Longitudinal Sleep Disruption in Shift-Working Healthcare Professionals: A Pilot Study

**DOI:** 10.1101/2025.09.19.25336161

**Authors:** Mohammed F. Salahuddin, Karn Sukararuji, Mahsa Sharifi, Kingsley Anetor Francis Odia, Md Dilshad Manzar, Seithikurippu R. Pandi-Perumal, Ahmed S. BaHammam

**Affiliations:** Department of Pharmaceutical Sciences, School of Pharmacy & Health Professions, Notre Dame of Maryland University, Baltimore, MD 21210, USA; Department of Nursing, College of Applied Medical Sciences, Majmaah University, Majmaah 11952, Saudi Arabia; Centre for Research and Development, Chandigarh University, Mohali 140413, Punjab, India; Division of Research and Development, Lovely Professional University, Phagwara 144411, Punjab, India; University Sleep Disorders Center, Department of Medicine, College of Medicine, King Saud University, Riyadh, Saudi Arabia; National Plan for Science and Technology, College of Medicine, King Saud University, Riyadh, Saudi Arabia

**Author notes:** Both authors have equally contributed to the work (Karn Sukararuji, Mahsa Sharifi). Corresponding Author Mohammed F Salahuddin, Department of Pharmaceutical Sciences, School of Pharmacy & Health Professions, Notre Dame of Maryland University, Baltimore, MD 21210, USA.

**Keywords:** Shift Work Schedule, Cortisol, Sleep Wake Disorders, Stress, Psychological, Sleep Diary

## Abstract

**Background:** Shift work is a well-established disruptor of sleep, yet the biological mechanisms driving sleep disturbances remain poorly understood. Salivary cortisol (HPA axis), α-amylase (sympathetic– adrenomedullary output), and DHEA-S (adrenal androgen with anti-glucocorticoid/resilience properties) are candidate indicators of stress-related sleep disruption. We therefore examined whether changes in these biomarkers were associated with 6-month sleep trajectories in health professionals.

**Methods:** In a prospective 6-month repeated-measures design, 52 healthcare professionals (daytime vs. rotating shifts; mean age 31.4 ± 9.4 years; 57% female) completed validated sleep assessments, PROMIS Sleep Disturbance, PROMIS Sleep Impairment, the Sleep-Wake Disorder Index (SWDI), and the NIH 7-day Sleep Diary, at baseline and six-month follow-up. Salivary cortisol, DHEA-S, and alpha-amylase were collected on the morning of Day 7 of each diary period. Change scores (Δ = follow-up – baseline) were computed. Repeated-measures ANOVA, Pearson correlations, and multivariable regressions assessed group differences and biomarker–sleep associations.

**Results:** Compared with daytime workers, rotating shift workers reported significantly greater increases in sleep disturbance, impairment, and reduced sleep efficiency over time (all p < 0.05). Reductions in cortisol and alpha-amylase were significantly associated with worsening PROMIS Sleep Disturbance and SWDI scores (r = –0.65 and –0.53, respectively; p < 0.05). Multivariable regression showed that decreased cortisol (β = −41.845, p = 0.0064) and increased DHEA-S (β = 0.001, p = 0.0405) associated with worsening PROMIS Sleep Impairment. A combined model including reduced cortisol, and increased DHEA-S associated with greater PROMIS Sleep Disturbance (adjusted R² = 0.698).

**Conclusion:** In this pilot, changes in salivary cortisol and DHEA-S were associated with longitudinal changes in sleep. These results suggest potential utility for biomarker-informed risk stratification, warranting confirmation in larger, controlled studies.

**Plain Language Summary:** People working rotating shifts often experience disrupted sleep and chronic stress due to irregular work hours. This study followed a group of rotating shift workers over six months to explore how stress levels, measured through saliva samples, and sleep quality changed over time. Researchers collected data on key stress hormones, including cortisol, alpha-amylase, and DHEA-S, and compared them with both subjective sleep surveys and daily sleep diaries.

The results showed that as stress biomarkers declined, participants reported worse sleep quality and greater impairment in daily functioning. This suggests that when the body’s stress response system becomes weakened, what scientists call “lower cortisol levels” it may be a sign of long-term health strain. These physiological changes mirrored growing sleep disturbances over time.

The findings highlight the importance of monitoring both sleep and biological stress markers in shift workers. Interventions that target better sleep hygiene and stress management may help protect the health of these essential workers. Future studies with larger, more diverse groups are needed to confirm these results and develop personalized strategies to reduce the negative effects of shift work.

**Graphical Abstract:** 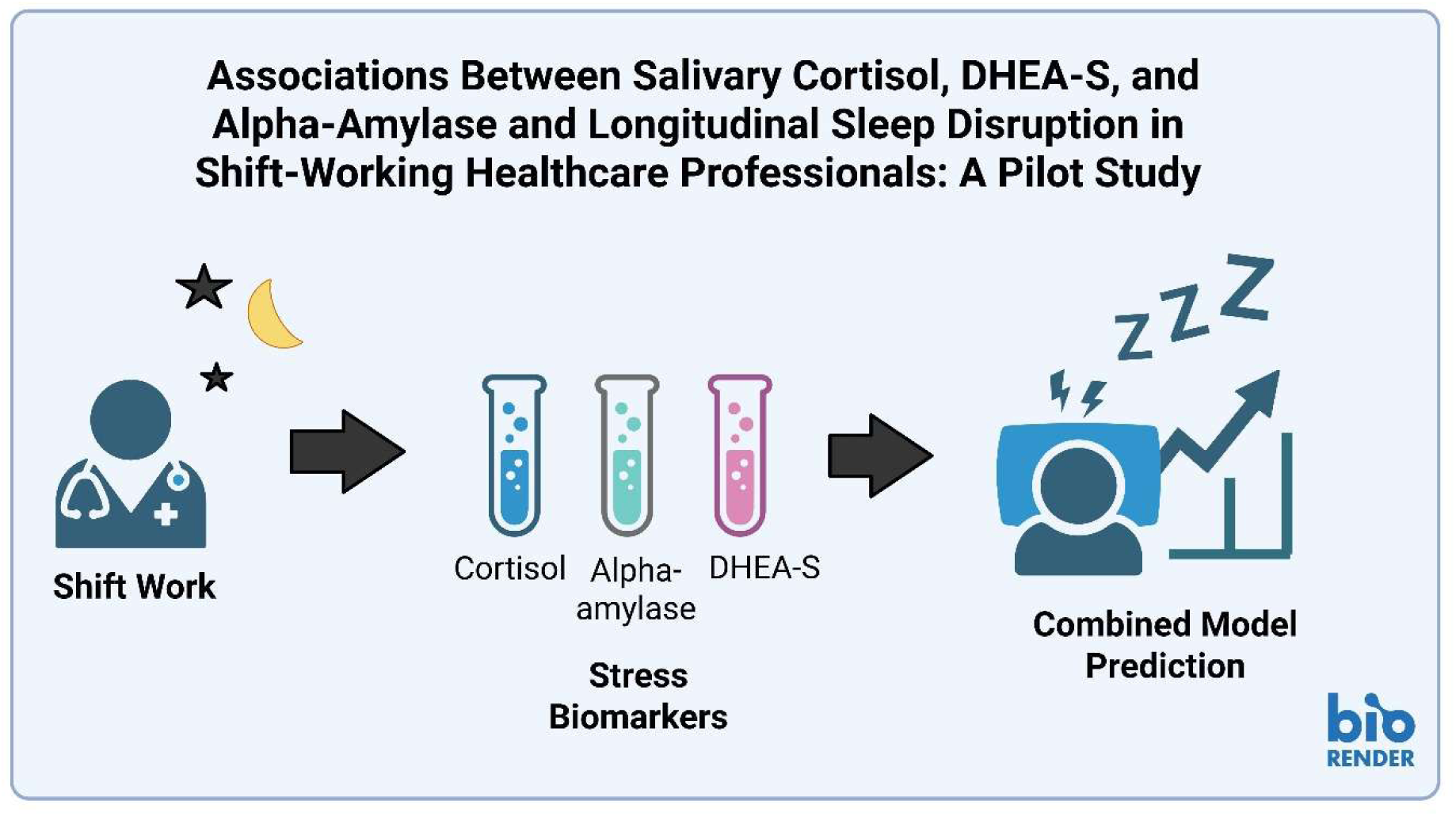

## 1. Introduction

Shift work, particularly rotating and night shifts, is highly prevalent among healthcare professionals and is strongly linked to inconsistent shift schedule, dysregulation of circadian timing system, forced sleep/wake misalignment, and elevated physiological and emotional distress ^1^. Emerging evidence highlights a bi-directional relationship between chronic stress and sleep, wherein dysregulation of the hypothalamic–pituitary–adrenal (HPA) axis contributes to insomnia, reduced sleep efficiency, and circadian rhythm disorders^2,3^. In shift-working populations, these interactions are compounded by light-at-night exposure, social jet lag, and irregular behavioral patterns^4,5^. Despite these known risks, longitudinal studies that integrate both subjective and objective sleep assessments with biological stress markers remain limited. Addressing this gap is essential for informing targeted interventions to reduce sleep-related morbidity in high-risk occupational populations.

Cortisol reflects HPA-axis activity with a diurnal rhythm; lower morning cortisol levels over time can accompany chronic stress and circadian strain^6^. Salivary α-amylase (sAA) is a noninvasive marker of sympathetic–adrenomedullary activation relevant to arousal and sleep continuity^7^. DHEA-S is an adrenal androgen with anti-glucocorticoid and neuroprotective actions; higher DHEA-S or a more favorable DHEA-S:cortisol balance has been proposed as an index of stress resilience^8^. Systematic and integrative reviews connect these biomarkers with cardiometabolic and sleep-related outcomes^9,10^ and with inflammatory/cellular pathways perturbed by short or disrupted sleep^11^. Despite this, few longitudinal studies in healthcare shift workers evaluate whether within-person biomarker changes track sleep deterioration across months, an evidence gap we address here.

Rotating shift workers have more than just "less sleep"; they also have numerous overlapping sleep issues that worsen over time, including circadian disruption, shift work sleep disorder (SWSD), chronic and persistent insomnia, excessive daytime sleepiness (EDS), and obstructive sleep apnea (OSA)^1^. This compounded health problems observed in rotating shift workers are caused by a long-standing conflict between artificial work schedules and the body’s evolutionary biology^1,4,5^. This issue transcends just "poor sleep"; it represents a systemic failure resulting from persistent circadian misalignment^1,4,5^.

Among UK night shift workers, approximately 24.5% meet criteria for shift work sleep disorder^12^. These disruptions are associated with significant cognitive and health impairments, underscoring the need for early identification and intervention^13^. Chronic exposure to shift work increases the risk of cardiometabolic and psychological disorders, largely mediated by sleep-wake misalignment and impaired physiological stress adaptation^14,15^. While prior studies have documented elevated perceived stress and poor sleep among shift workers^16,17^, longitudinal studies involving biological markers of stress in healthcare professionals remain scarce. Across prospective longitudinal work, salivary α-amylase (sAA) tracks stress more consistently than cortisol, and the sAA: cortisol ratio captures sustained sympathetic-HPA asymmetry in teachers, higher ratios prospectively predicted greater perceived stress and psychosomatic strain despite null cross-sectional cortisol effects^7^. Experimental training settings exhibit a similar pattern, with sAA rising under acute demands, while cortisol often remains unchanged, indicating that sAA may be a more sensitive proximal index of stress load^6^. Beyond symptoms, both cortisol and sAA relate to cardiometabolic risk, underscoring their clinical relevance^9^. Building on this literature, we prospectively test whether 6-month changes in cortisol, DHEA-S (an anabolic/adrenocortical counter-regulator), and sAA and their ratios, are associated with concurrent changes in sleep disturbance and impairment among shift-working healthcare professionals.

Salivary biomarkers offer non-invasive, repeatable indicators of stress system activation. Cortisol reflects HPA axis function and is sensitive to circadian disruption^18^. Alpha-amylase represents sympathetic nervous system activity and correlates with acute stress and sleep loss^19,20^. Dehydroepiandrosterone sulfate (DHEA-S), an anabolic hormone, contributes to resilience and its ratio to cortisol may index adaptive stress-buffering capacity^8^. While cross-sectional studies have shown altered biomarker levels in shift workers^21^, there is a lack of longitudinal data linking these physiological changes to evolving sleep outcomes^22^. This study builds upon our earlier cross-sectional analysis of the same cohort, which focused on the mediating role of negative mood affect in the perceived stress–insomnia relationship among student pharmacist shift workers^23^. In contrast, the current longitudinal study incorporates salivary biomarkers and repeated sleep assessments to explore physiological stress trajectories and their predictive role in sleep disruption over time, providing novel insights into temporal dynamics of stress adaptation.

To address this gap, we conducted a six-month prospective study assessing associations between salivary biomarkers and sleep outcomes in healthcare professionals working rotating and daytime shifts. We hypothesized that blunted cortisol and DHEA-S trajectories would predict worsening sleep. Our goal was to identify early physiological signatures of stress-related sleep impairment in shift workers to support future biomarker-informed interventions.

## 2. Materials and Methods

### 2.1. Study Design and Participants

This prospective observational study enrolled 52 healthcare professionals from Notre Dame of Maryland University. Participants were classified as either: (1) rotating shift workers (schedules including hours past 9:00 PM or before 6:00 AM) or (2) regular daytime workers (9:00 AM–5:00 PM). Inclusion criteria were: (a) employment ≥1 year in a clinical setting, (b) age 18–60 years, and (c) willingness to complete a 7-day sleep diary and provide salivary samples at baseline and six-month follow-up. Exclusion criteria included pregnancy, psychiatric disorders, chronic sleep-impairing conditions (e.g., sleep apnea, chronic pain), and use of corticosteroids, hormone therapy or psychotropic medications within 30 days, long-haul (≥3time zones) air travel in the prior 4 weeks, known to influence HPA axis function.

The institutional review board (IRB) approval was obtained from Notre Dame of Maryland University (Approval No: IRB # PH053023MSDS), and written informed consent was secured from all participants prior to enrollment. Data collection occurred in two phases, baseline and six-month follow-up, to observe within-subject physiological adaptation while minimizing attrition in this high-demand workforce.

#### 2.1.1. Shift Work Composite Score

To quantify cumulative circadian strain, a Shift Work Composite Score (range: 4–13) was constructed from four ordinal components:

- Shift type (1 = daytime, 2 = rotating/night)
- Shift duration (1 = ≤8h, 2 = 8–12h, 3 = >12h)
- Tenure on shift (1 = 1–3 months to 5 = >24 months)
- Weekly hours (1 = 20h, 2 = 40h, 3 = >40h)

Each element reflected work-related circadian disruption per prior studies^17,24,25^. The additive score captured cumulative shift burden without normalizing ordinal components.

### 2.2. Sleep Assessment

Participants completed the NIH Consensus Sleep Diary for seven days at each timepoint^27^. This approach offers validated week-long estimates of sleep patterns with high reliability (ICC ≥ 0.60)^27^. On Day 7 of each assessment week, participants completed:

- PROMIS Sleep Impairment Scale v1.0–SF 8a (functional impact of poor sleep)^27^
- PROMIS Sleep Disturbance Scale v1.0–SF 8b (difficulty falling/staying asleep) ^27^
- Shift Work Disorder Index (SWDI) (symptoms of insomnia, daytime dysfunction)^28^ Diary-derived (self-reported) metrics included:
- Total Sleep Time (TST)
- Wake After Sleep Onset (WASO)
- Sleep Efficiency (SE%) = TST ÷ time in bed

Internal consistency was PROMIS Sleep Impairment (0.80), Sleep Disturbance (0.70), and SWDI (0.90) indicated good to excellent internal consistency in this sample.

Δ (change) scores were computed as: **Δ** = Follow-up (Time 2) – Baseline (Time 1) Positive Δ values reflected worsening sleep outcomes.

### 2.3. Salivary Biomarker Collection and Analysis

Saliva was collected via passive drool on the morning of Day 7 at ∼9:00 AM at both timepoints. Participants followed strict protocols: (1) no food/drink (except water), brushing, smoking for 30 minutes; (2) no alcohol within 12 hours; (3) recorded caffeine, medication, and menstrual cycle phase; (4) no strenuous activity before collection. Samples were frozen at −80°C until assay.

Biomarkers analyzed using duplicate ELISA (Salimetrics, Carlsbad, CA) included:

- Cortisol (HPA axis marker)
- Alpha-amylase (sympathetic activity marker)
- DHEA-S (stress resilience marker)

Plates were read at 450 nm on a CLARIOstar Plus microplate reader (BMG Labtech, Cary, NC). All assays had CVs <10%, indicating excellent precision. Δ biomarker scores (Follow-up − Baseline) were calculated to assess longitudinal physiological changes.

### 2.4. Statistical Analysis

Data were analyzed using StatView (Version 5.0.1, SAS Institute). Normality was assessed via the Shapiro-Wilk test. All variables met assumptions (p > 0.05) except baseline SWDI (p = 0.002) and Δ sleep disturbance (p = 0.0098). Given the robustness of ANOVA and Pearson correlation to modest non-normality in pilot studies, parametric methods were used without transformation. Levene’s test confirmed homogeneity of variance, except for 6-month PROMIS Sleep Impairment (p = 0.030); assumptions were deemed acceptable. A post hoc power analysis (G*Power 3.1) indicated >90% power to detect large within-subject effects (f = 0.40, n = 52). Biomarker correlations (n = 14) retained >80% power for large effects (r ≥ 0.60).

Repeated-measures ANOVA evaluated within-subject changes in subjective (PROMIS, SWDI) and diary derived (TST, SE%) sleep metrics over time. Between-subject factor: shift type. Time × Shift Type interactions were nonsignificant and are not reported.

Pearson correlations (N = 14) assessed relationships between Δ biomarkers and Δ sleep outcomes. Multivariable linear regression models examined the independent predictive contributions of Δ cortisol and Δ DHEA-S to PROMIS Sleep Impairment and Disturbance scores. For each outcome we removed a small number of physiologically implausible values (TST = 1; sleep efficiency = 2; sleep disturbance = 1), fitted a Group × Time model comparing change between shifts, and repeated the analysis with the full dataset and after influence screening using established diagnostic criteria (|studentized residual| > 3, Cook’s D > 4/n, leverage > 2p/n, DFFITS > 2√(p/n)). Results for all three specifications are shown in Supplementary Table S1. For salivary biomarker analysis, only a subset of participants (N = 14) who provided both baseline and follow-up samples were included (Supplementary Table S2). For sleep diary and survey measures, StatView automatically excludes participants with incomplete paired data, ensuring only complete datasets are used in repeated measures analysis. This approach aligns with ethical data handling practices, avoiding imputation in a pilot setting. All tests were two-tailed, with p ≤ 0.05 considered significant. To address potential inflation of type I error from multiple comparisons, Benjamini–Hochberg FDR correction (α_FDR = 0.05) was applied to the biomarker and sleep outcome analyses; raw and adjusted p-values are provided in Supplementary Table S3. Confidence intervals for regression coefficients were calculated using the standard formula (β ± 1.96 × SE) and are presented as point estimates to aid interpretation, without formal inferential generalization due to the pilot sample size (N = 14). Effect sizes included partial η² for ANOVA and standardized β and R² for regression.

This was a two-timepoint prospective repeated-measures pilot (baseline and 6-month follow-up). Because there are exactly two timepoints, the between-group difference in change (Follow-up − Baseline) is reported alongside the RM-ANOVA as the two-timepoint analogue of the Shift×Time term. To reduce overfitting in the biomarker subsample, regression models were pre-specified and parsimonious (two predictors: Δcortisol, ΔDHEA-S; per-model N=14 complete cases; no stepwise selection; VIF<2). For transparency, primary results are accompanied by sensitivity checks (with outliers included and after influence screening), with Ns and p-values summarized in Supplementary Table S1. As detectable-effect context, with n=14 and k=2 predictors (α=0.05) the design affords ∼80% power only for large effects (f²=0.44; R²=0.31); findings are interpreted as exploratory.

## 3. Results

### 3.1. Sociodemographic and Sleep Characteristics

The sample consisted of 52 healthcare professionals (mean age: 31.37 ± 9.37 years), with a slightly higher proportion of females (57%) than males (42.3%). At baseline, groups were broadly comparable; men in rotating shifts were somewhat older on average than men in day shifts (37.4 ± 10.4 vs 32.1 ± 9.0 years; Regular n=13 men/Rotating n=8 men), whereas women were similar in age across groups (29.1–29.7 years; Regular n=23 women/Rotating n=8 women) (see Supplementary Table S4). Most participants resided in urban areas (84.6%) and had a bachelor’s or doctoral degree. The ethnic distribution was diverse, and the average Shift Work Composite Score was 8.04 ± 2.74. Income levels varied, with the largest group earning above $100,000 (25%). At baseline, average PROMIS Sleep Impairment and Sleep Disturbance T-scores were 57.25 and 53.02, respectively, indicating elevated sleep-related concerns. Diary derived sleep measures revealed an average total sleep time of ∼433 minutes (7.2 hours), with relatively short sleep latency (21.4 min) and low wake after sleep onset (6.9 min), suggesting some preserved aspects of sleep continuity despite subjective impairments (Table 1).

**Table 1.**
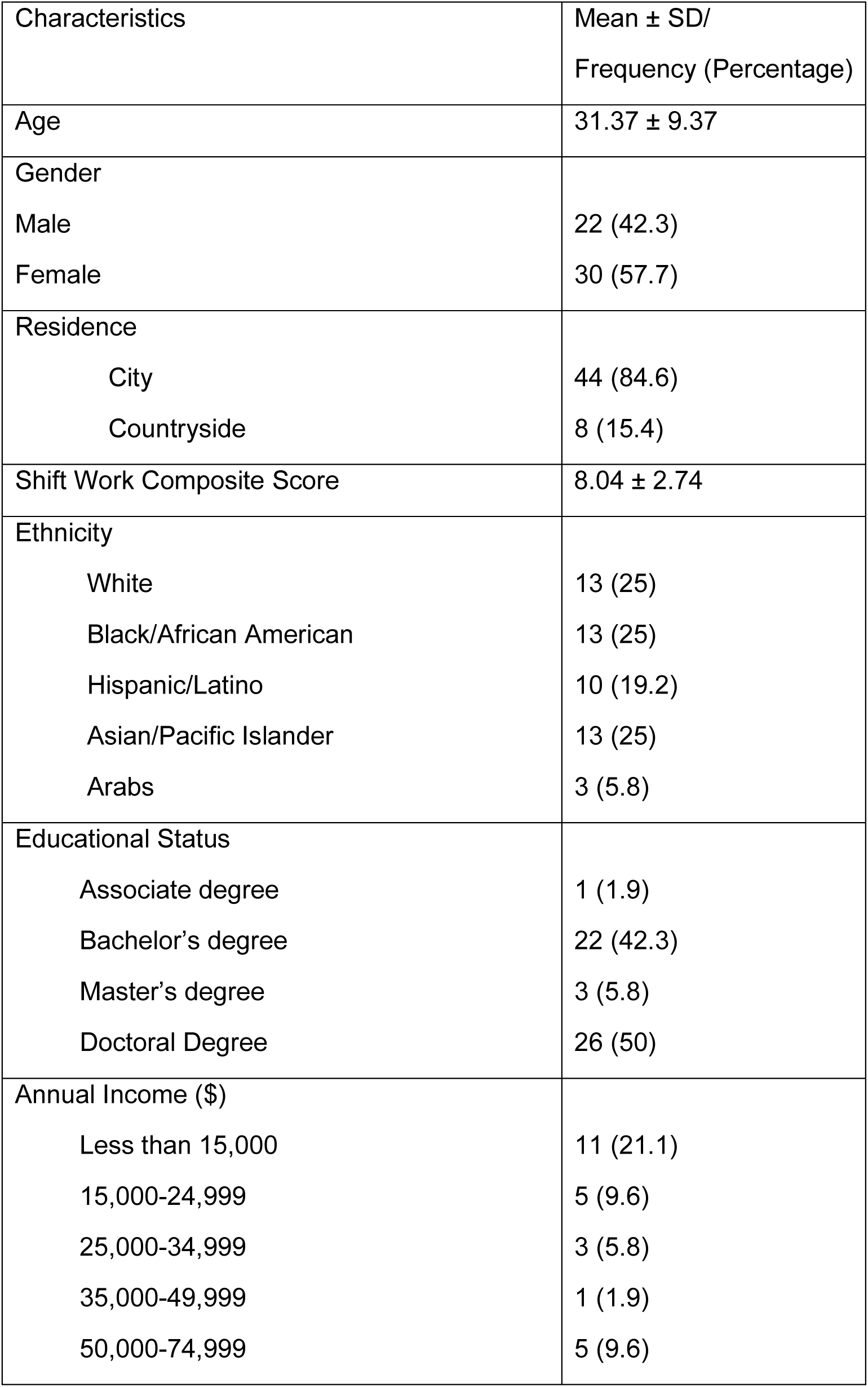
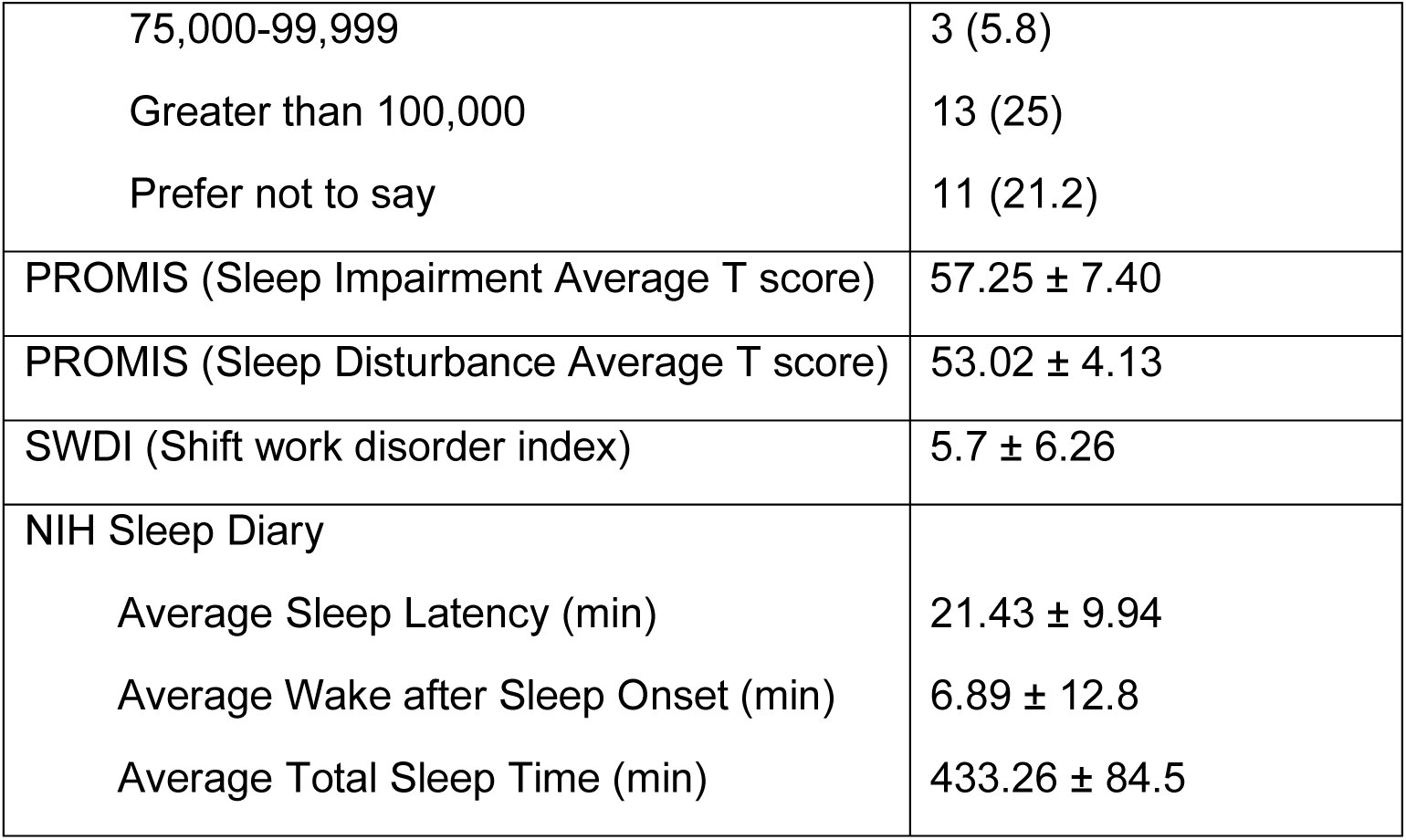
Sociodemographic characteristics of healthcare professionals. Data are presented as Mean ± SD or Frequency (Percentage).

### 3.2. Shift Work and Sleep Outcomes

Group comparisons for sleep outcomes use repeated-measures ANOVA by shift type; biomarker–sleep analyses include all participants with paired saliva (n = 18). Rotating shift workers showed significant longitudinal increases in sleep impairment, indicated by higher PROMIS sleep disturbance (*F*(1,36)=6.47, *p*=0.0154, *FDR p* =0.0330, η² = 0.15; Fig 1A), PROMIS Sleep Impairment scores (*F*(1,36)=5.94, *p*=0.0198, *FDR p*=0.03713, η² = 0.14; Fig 1B), SWDI scores (F(1,37)=29.68, *p*<0.0001, *FDR p*= 0.0015, η² = 0.45; Fig 1C) relative to daytime workers.

**Figure 1.**
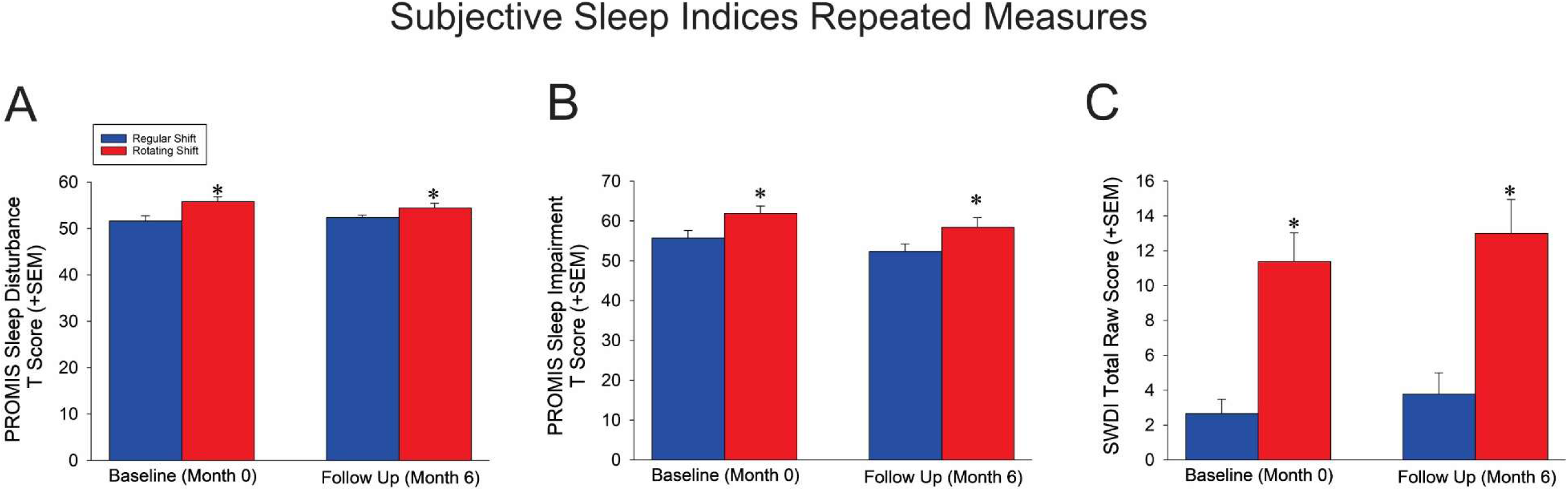
Subjective sleep outcomes across two time points (Baseline and 6-Month Follow-Up) among regular healthcare workers. (A) PROMIS Sleep Disturbance T-scores; (B) PROMIS Sleep Impairment T-scores; and (C) Shift Work Disorde scores. Across all three measures, rotating shift workers reported significantly greater sleep disturbance, impair related symptoms compared to regular day-shift workers, both at baseline and follow-up. Data are presented as 0.05 for group comparison between rotating and regular shifts.

Diary derived sleep measures also trended toward impairment in rotating shift workers, notably decreased total sleep time (*F*(1,12)=4.75, *p*=0.05, *FDR p*= 0.05, η² = 0.28; Fig 2A) and sleep efficiency (*F*(1,11)=5.33, *p*=0.0414, *FDR p*= 0.04777, η² = 0.33; Fig 2B).

**Figure 2.**
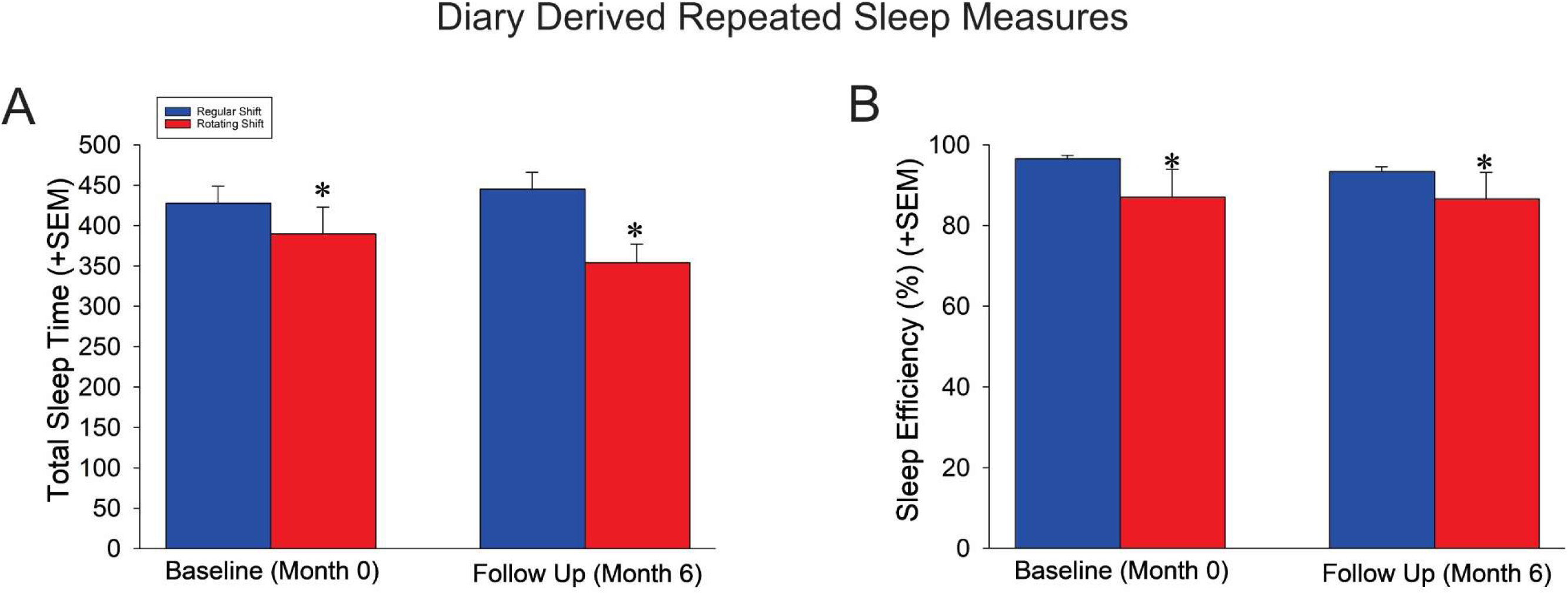
Diary derived (self-reported) sleep outcomes across two time points (Baseline and 6-Month Follow-U rotating shift healthcare workers. (A) Total Sleep Time (minutes); (B) Sleep Efficiency (%). Rotating shift workers consistently exhibited shorter to lower sleep efficiency compared to regular shift workers at both baseline and follow-up. Data are presented a 0.05 for group comparison between rotating and regular shifts.

### 3.3. Salivary Biomarkers and Sleep Associations

In this longitudinal study, we evaluated how changes in salivary stress biomarkers from baseline (Day 7 of the first timepoint) to follow-up (Day 7 of the six-month timepoint) were associated with concurrent changes in subjective sleep health among shift working healthcare professionals. All delta values were calculated as follow-up minus baseline, such that negative values represent a decline over time and positive values reflect an increase.

#### Standalone Cortisol

A significant negative association was observed between changes in salivary cortisol and PROMIS Sleep Disturbance scores (*β* = –22.29, *r* = –0.653, *p* = 0.0113, *FDR p*= 0.0305, *R²* = 0.427; Fig. 3A), indicating that lower cortisol over time were associated with greater self-reported disturbance in sleep, these cross-sectional associations do not establish causality A similar pattern was seen for PROMIS Sleep Impairment scores (*β* = – 36.66, *r* = –0.594, *p* = 0.0251, *FDR p*= 0.0377, *R²* = 0.353; Fig. 3B), suggesting that HPA-axis downregulation may exacerbate vulnerability to poor sleep quality.

**Fig 3.**
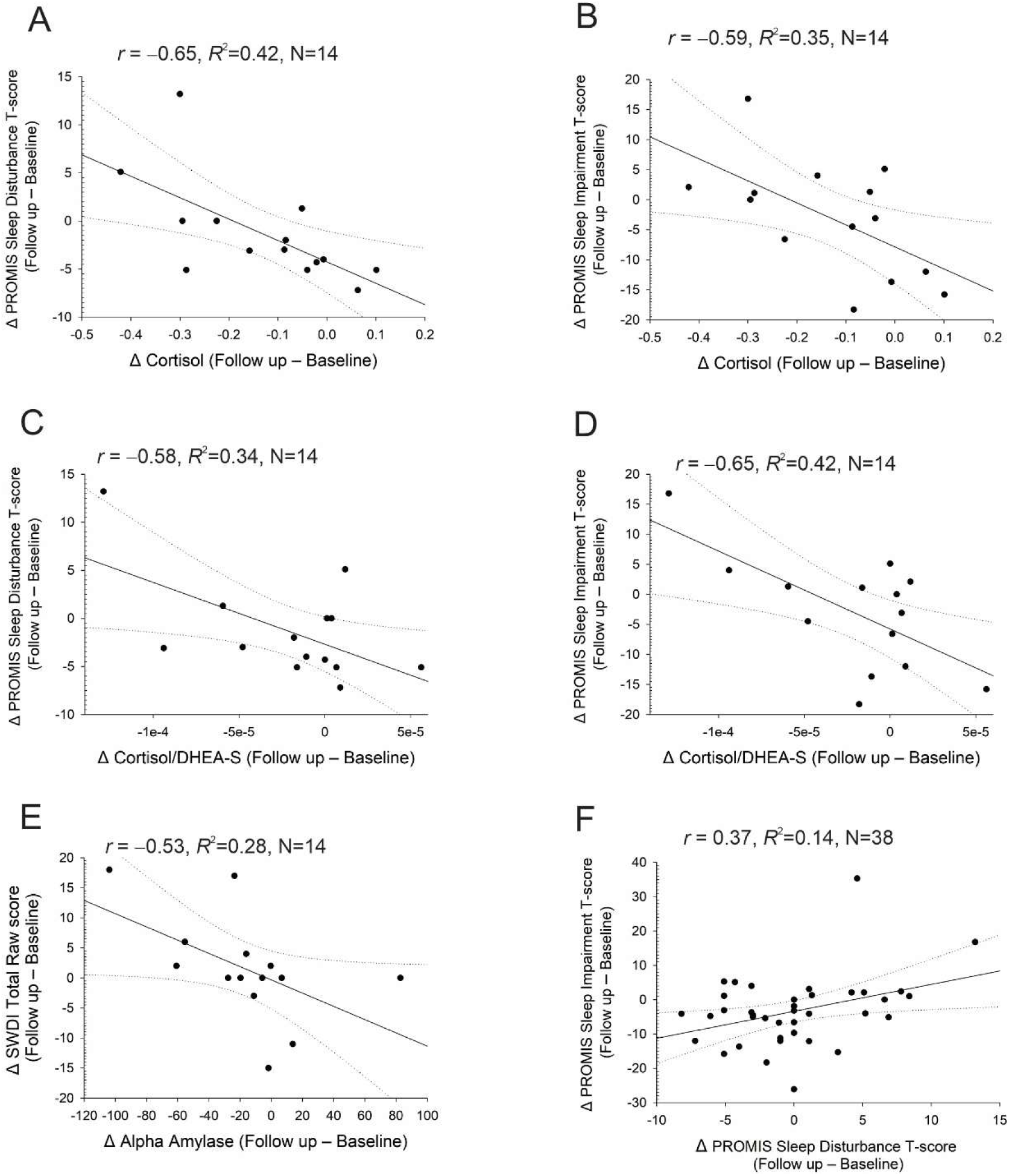
Correlations between changes in salivary biomarkers and subjective sleep outcomes from baseline to 6-month follow-up. (A–B) Greater decreases in salivary cortisol levels were significantly associated with worsening PROMIS Sleep Disturbance (A) and PROMIS Sleep Impairment (B) T-scores. (C–D) Declines in the cortisol-to-DHEA-S ratio were also associated with greater sleep disturbance (C) and impairment (D). (E) Reductions in salivary alpha-amylase were significantly correlated with increased SWDI scores, reflecting worsening subjective sleep quality. (F) Changes in PROMIS Sleep Disturbance scores were moderately associated with changes in Sleep Impairment scores. All plots show standardized residuals with 95% confidence bands. Negative Δ values indicate biomarkers decrease over time. Pearson’s r and R² are shown. Analyses are exploratory. Panel Ns: 3A = 14, 3B = 14, 3C = 14, 3D = 14, 3E = 14 (biomarker subsample, complete cases); 3F = 38 (sleep-scale)

#### Cortisol/DHEA-S Ratio (C/D)

The ratio of cortisol to DHEA-S, a marker of anabolic-catabolic balance, was inversely associated with changes in both Sleep Disturbance (*β* = –64,337.54, *r* = –0.581, *p* = 0.0292, *FDR p*=0.0398, *R²* = 0.338; Fig. 3C) and Sleep Impairment (*β* = – 129,675.56, *r* = –0.648, *p* = 0.0122, *FDR p*= 0.0305, *R²* = 0.420; Fig. 3D). These results are consistent with the idea that a lower cortisol:DHEA-S balance is associated with worse sleep; directionality cannot be inferred.

#### Alpha-Amylase

Declines in salivary alpha-amylase, a marker of sympathetic nervous system activity, were significantly associated with increases in Sleep-Wake Disorder Index (SWDI) scores (*β* = –0.110, *r* = –0.532, *p* ≤ 0.05, *FDR p*=0.05, *R²* = 0.283; Fig. 3E), supporting the notion that reduced sympathetic arousal may accompany deterioration in circadian regulation.

#### Sleep Domain Correlation

PROMIS Sleep Disturbance and Sleep Impairment scores were positively correlated over time (*β* = 0.785, *r* = 0.369, *p* = 0.0224, *FDR p*= 0.03733, *R²* = 0.137; Fig. 3F), suggesting overlapping disruption across multiple dimensions of sleep.

Because these correlations derive from a small biomarker subsample and two timepoints, they are exploratory and R² values should be interpreted cautiously.

### 3.4. Multivariable Predictors of Sleep Impairment

To further examine the joint influence of biological and behavioral factors on sleep health, multivariable linear regression models were performed. In the model predicting changes in PROMIS Sleep Disturbance T-scores, significant predictors included salivary cortisol (β = – 25.792, 95% CI [–36.16, –15.42], *p* = 0.0005, *FDR p*=0.0038), and salivary DHEA-S (β = 0.001, 95% CI [0.00056, 0.00144], *p* = 0.0035, *FDR p*= 0.0175; Table 2), with an Adjusted R² of 0.698.

**Table 2.** Merged Multivariable Regression Analysis of Predictors.

| Variable | Coefficient | SE | 95% CI (low) | 95% CI (high) | P-Value | Model |
| --- | --- | --- | --- | --- | --- | --- |
| Δ Salivary Cortisol ( Follow up — Baseline) | -25.792 | 5.29 | -36.16 | -15.424 | 0.0005 | Δ PROMIS Sleep Disturbance T score (Follow up — Baseline) |
| Δ Salivary DHEA-S ( Follow up — Baseline) | 0.001 | 0.0002 | 0.0006 | 0.00144 | 0.0035 |  |
| Δ Salivary Cortisol ( Follow up — Baseline) | -41.845 | 12.469 | -66.284 | -17.406 | 0.0064 | Δ PROMIS Sleep Impairment T score (Follow up — Baseline) |
| Δ Salivary DHEA-S ( Follow up — Baseline) | 0.001 | 0.0004 | 0.00016 | 0.0018 | 0.0405 |  |

**Notes: SE – standard error; CI – confidence interval; DHEA-S – dehydroepiandrosterone sulfate**

Notably, lower cortisol and higher DHEA-S levels were independently associated with greater increases in sleep disturbance, suggesting dysregulated stress response and reduced anabolic support as contributing factors.

In a separate model predicting changes in PROMIS Sleep Impairment T-scores, both salivary cortisol (β = –41.845, CI [–66.28, –17.41], *p* = 0.0064, *FDR p*=0.0240) and DHEA-S (β = 0.001, CI [–0.00096, 0.00296], *p* = 0.0405, *FDR p*= 0.04777; Table 2) were independently associated, consistent with altered stress-system balance; however, these associations are not evidence of causation. (Adjusted R² = 0.487).. Confidence intervals are reported as point estimates to aid interpretation, recognizing the exploratory nature of this pilot analysis.

To account for multiple comparisons, Benjamini-Hochberg FDR corrections were applied. All significant findings remained robust after correction (see Supplementary Table S3).

## 4. Discussion

This longitudinal study explored the relationship between physiological stress biomarkers and sleep outcomes in healthcare professionals, highlighting the vulnerability of rotating shift workers. Notably, decreases in salivary cortisol, alpha-amylase, and DHEA-S over six months were significantly associated with worsening subjective and diary derived sleep measures, suggesting that blunted stress system output may indicate allostatic overload and emerging sleep pathology.

### Shift Work and Sleep Vulnerability

Rotating shift workers exhibited greater sleep impairment over time, with PROMIS and SWDI scores reflecting persistent subjective disturbance. Nearly half the variance in SWDI scores (η² = 0.45) was attributed to shift type, aligning with prior findings that irregular schedules increase circadian misalignment and chronic fatigue^23,29^. These disruptions not only affect personal wellbeing but may impair clinical performance and safety^30^. Tailored fatigue mitigation strategies, including chronotherapy and biomarker monitoring, are essential to reduce occupational health risks.

We considered experience/tenure as a potential modifier; however, in this pilot the tenure cells were too small to model reliably. Notably, evidence for durable ‘acclimation’ is mixed, and large cohort/field studies suggest that longer rotating night-shift tenure is associated with worse long-term health and that recommending permanent nights to improve sleep is not supported^31,32^.

### Individual differences and rhythm recovery

Susceptibility to shift-related sleep disruption likely varies by sex and other traits (e.g., chronotype, tenure), with heterogeneous recovery after schedule changes. Beyond sleep hygiene, circadian alignment strategies, including timed light exposure and schedule regularization, are associated with better cardiometabolic profiles in population-scale data and offer plausible pathways for rhythm ‘recovery’ in shift workers^33^. These considerations suggest our estimates may mask subgroup effects that future work should test explicitly.

### Lower Morning Cortisol and Stress-Sleep Disruption

Greater declines in cortisol were significantly associated with worsening sleep disturbance (r = – 0.65, R² = 0.42) and impairment (r = –0.59, R² = 0.35), consistent with lower morning cortisol levels as a maladaptive response to chronic stress and circadian strain ^34,35^. Rather than reflecting recovery, reduced cortisol may indicate HPA axis exhaustion, diminished physiological flexibility, and impaired arousal regulation^36^. This mirrors patterns seen in burnout, PTSD, and chronic fatigue.

Although DHEA-S is often discussed as stress-buffering, empirical links to sleep are heterogeneous across populations, biospecimens, and analytic contexts. DHEA/DHEA-S can increase with stress exposure as a short-term, compensatory response, a pattern reported in experimental and occupational studies, which may help explain positive associations with sleep disturbance in some settings; by contrast, diurnal DHEA rhythms sometimes show weak or inconsistent relations to global sleep quality^37,38^. In our pilot, the absolute DHEA-S signal versus its balance with cortisol may capture different facets of adaptation vs. strain; larger studies should test non-linear and ratio-based models.

Similarly, alpha-amylase decline correlated with increased SWDI scores (r = –0.53, R² = 0.28), suggesting blunted sympathetic activity may also reflect diminished stress adaptability^39,40^. These findings validate the use of both subjective scales and objective biomarkers in detecting early physiological disruption.

### Implications for intervention design

These findings offer a foundation for integrating biomarker monitoring into occupational health programs targeting sleep health. Future interventional studies could evaluate whether improvements in sleep hygiene, schedule stability, or behavioral therapies (e.g., CBT-I, mindfulness-based stress reduction) are associated with normalization of stress biomarkers and improved sleep outcomes. Evidence-based CBT-I (sleep consolidation/restriction, stimulus control, cognitive restructuring, sleep scheduling) improves insomnia outcomes across populations and is feasible to adapt for shift workers and digital delivery^41^. In parallel, circadian schedule/light interventions (e.g., strategic light exposure/avoidance and stabilized rotations) warrant testing alongside sleep skills training. Lifestyle levels are also relevant: higher physical activity is associated with fewer sleep disturbances, partly via inflammatory pathways^42^, and reducing environmental pollutant exposure (e.g., cadmium) is linked to lower sleep-disturbance risk, with exercise potentially mitigating pollutant-related sleep harm^43^. These literatures motivate multicomponent trials tailored to shift-work constraints. Larger, multicenter trials incorporating actigraphy and cortisol awakening response (CAR) measures would enhance the precision of physiological tracking^44^. Future research should (i) increase sampling frequency (multi-day biomarker panels to model within-person dynamics) and add objective sleep/light (7– 14 days actigraphy with integrated light sensors); (ii) include cortisol-awakening response (CAR) to index HPA reactivity; (iii) pre-register analyses and power for sex-stratified and chronotype-stratified effects; and (iv) randomize schedule/light modules and/or CBT-I components to estimate additive vs. synergistic benefits. Finally, sex-specific analyses and exploration of resilience pathways (e.g., DHEA-S modulation) could support more personalized, preventative strategies to improve sleep and reduce burnout in chronically strained healthcare workforces.

Multivariable models reinforced the importance of cortisol and DHEA-S: both independently associated with sleep disturbance (Adjusted R² = 0.698) and impairment (R² = 0.487). DHEA-S decline was linked to reduced neuroendocrine resilience and poorer sleep, aligning with its known role in stress buffering^45,46^.

### Limitations

This study’s modest sample size (n = 52) limits generalizability, and only two timepoints constrain dynamic modeling. No formal power analysis was conducted due to the pilot sample size and exploratory design. While validated subjective tools were used, diary derived sleep metrics were limited to diaries; actigraphy and polysomnography were not feasible but represent important future directions. An additional limitation is that cortisol was measured only once in the morning at each timepoint, which may not capture diurnal or day-to-day variability and could affect the interpretation of HPA axis activity. Furthermore, diurnal variation, caffeine intake, seasonal factors, and menstrual cycle phase were not fully controlled, all of which could influence biomarker levels and sleep outcomes.

## 5. Conclusion

Shift workers encounter a heightened risk of medical, psychological, and metabolic health complications. In this pilot cohort, six-month declines in salivary cortisol, α-amylase, and DHEA-S were associated with worsening sleep measures among healthcare professionals. While causal inference is not possible from this observational design, these associations may help inform risk stratification approaches and guide the design of adequately powered randomized studies testing whether CBT-I, circadian interventions, and lifestyle strategies can improve both sleep and biomarker profiles in shift-working clinicians.

## Acknowledgements

We are grateful to the participants of the study. While preparing this work, the author(s) used ChatGPT to polish the text. After using this tool/service, the author(s) reviewed and edited the content as needed and took full responsibility for the publication’s content. The graphical abstract was created in BioRender. Mohammed, S. (2025) https://BioRender.com/2wq609y

This is a preprint. It has been submitted for publication at Nature and Science of Sleep

## Ethical and Institutional Review Board

The present study was approved by the Notre Dame of Maryland University IRB board vide # PH053023MSDS. Consent was obtained by the study participants prior to study commencement. The authors assert that all procedures contributing to this work comply with the ethical standards of the relevant national and institutional committees on human experimentation and with the Helsinki Declaration of 1975, as revised in 2013.

## Data Availability Statement

The data is available at reasonable request to the corresponding author.

## Author Contribution Statement

**Mohammed F. Salahuddin:** Conceptualization; Methodology; Formal Analysis; Software; Visualization; Writing – Original Draft; Writing – Review & Editing; **Karn Sukararuji:** Data Curation; Investigation; Writing – Original Draft; Writing – Review & Editing; **Mahsa Sharifi:** Data Curation; Investigation; Writing – Original Draft; Writing – Review & Editing; **Kingsley Odia:** Formal Analysis; Writing – Original Draft; Writing – Review & Editing; **Md Dilshad Manzar:** Visualization; Formal Analysis; Validation; Writing – Original Draft; Writing – Review & Editing; **Seithikurippu R. Pandi-Perumal:** Conceptualization; Methodology; Supervision; Writing – Original Draft; Writing – Review & Editing; **Ahmed S. BaHammam:** Conceptualization; Methodology; Supervision; Writing – Original Draft; Writing – Review & Editing.

All authors contributed to the conception and design, as well as data acquisition/analysis/interpretation; drafted or critically revised the manuscript; approved the final version; agreed on the journal of submission; and agreed to be accountable for all aspects of the work.

